# Immune checkpoint inhibitor response in sarcomas associates with immune infiltrates and increased expression of transposable elements and viral response pathways

**DOI:** 10.1101/2024.01.02.24300710

**Authors:** Benjamin A. Nacev, Martina Bradic, Hyung Jun Woo, Allison L. Richards, Ciara M. Kelly, Mark A. Dickson, Mrinal M. Gounder, Mary L. Keohan, Ping Chi, Sujana Movva, Robert Maki, Emily K. Slotkin, Evan Rosenbaum, Viswatej Avutu, Jason E. Chan, Lauren Banks, Travis Adamson, Samuel Singer, Cristina R. Antonescu, William D. Tap, Mark T.A. Donoghue, Sandra P. D’Angelo

## Abstract

Response to immune checkpoint inhibition (ICI) in sarcoma is overall low and heterogeneous. Understanding determinants of ICI outcomes may improve efficacy and patient selection. One potential mechanism is epigenetic de-repression of transposable elements (TEs), which stimulates antitumor immunity. Here, we used transcriptomic data to assign immune-hot versus immune-cold status to 67 pre-treatment biopsies of sarcomas from patients treated on ICI trials. Progression-free survival and overall response was superior in the immune-hot group. Expression of TEs and epigenetic regulators significantly predicted immune-hot status in a regression model in which specific TE subfamilies and *IKZF1*, a chromatin-interacting transcription factor, were significantly contributory. TE and *IKZF1* expression positively correlated with tumor immune infiltrates, inflammatory pathways, and clinical outcomes. Key findings were confirmed in a validation cohort (n=190). This work suggests that TE and *IKZF1* expression warrant investigation as predictive biomarkers for ICI response and as therapeutic targets in sarcomas.

## INTRODUCTION

Sarcomas are a diverse group of more than 170 histologic entities^1^ that derive from tissues of mesenchymal origin. The underlying genetic causes of sarcomas^2,3^ are diverse and their biologic and pathologic behavior is highly varied^4^. The clinical management of metastatic sarcomas is generally palliative and relies upon systemic therapies including cytotoxic chemotherapy, targeted therapies, and in some instances immune checkpoint blockade^5^. The overall response rate to first-line chemotherapy in soft tissue sarcoma is approximately 20%^6^. Hence, there is a need for both new treatment modalities and improved methods to select patients most likely to respond to specific treatments.

Immune checkpoint inhibition (ICI) has been studied in sarcomas, and activity has been noted with nivolumab (anti-PD-1) alone or in combination with ipilimumab (anti-CTLA-4)^7^ or with pembrolizumab (anti-PD-1) as a single agent^8^. Efforts to enhance the activity of ICI through combination with other immune-modulatory drugs or with cytotoxic therapies has revealed variable response rates, which likely depend on the drug combination and sarcoma subtype (recently reviewed^9^). In parallel, predictive biomarkers for ICI response in sarcoma are being explored. While microsatellite instability (MSI) and high tumor mutation burden (TMB) predict response in carcinomas^10–16^, TMB is relatively low in sarcomas and MSI is exceedingly rare^17^. Alternative biomarkers such as tertiary lymphoid structures and B cell and CD8+ T cell infiltrates correlate with ICI response in some soft tissue sarcomas such as undifferentiated pleomorphic sarcoma (UPS)^2,18,19^.

Another potential determinant and predictor of antitumor immunity and ICI response are epigenetic states, which are determined by chemical modifications of DNA, RNA, and DNA-associated proteins together with their positioning relative to specific genomic sequences^20^. One key function of epigenetic states is to regulate transcriptional programs, including those that influence immune signaling. Therefore, genetic or pharmacologic perturbation of the machinery that establishes or maintains epigenetic states can prime ICI response in preclinical models and correlates with ICI clinical response^21–29^. For example, epigenetic mechanisms can promote immune escape through repression of antigen-presenting machinery and transposable elements (TEs), epigenetically silenced sequences of viral origin that, when de-repressed, stimulate antiviral immune signaling^22,24,30^.

We therefore hypothesized that sarcoma baseline immune infiltrates and clinical outcomes following immunotherapy treatment are influenced by expression of TEs and epigenetic regulators. To test this, we generated and analyzed transcriptomic profiles of pre-treatment biopsies from 67 unique patients enrolled in 3 ICI trials at our institution and an independent validation cohort. Here, we demonstrate that the efficacy of ICIs is linked to the de-repression of TEs that are normally silenced by epigenetic mechanisms and upregulation of the transcription factor IKZF1, which interacts with chromatin-modifying complexes. TE and *IKZF1* upregulation in turn correlate with hallmarks of tumor-intrinsic innate immune activation such as type I interferon and antigen presentation, suggesting a potential mechanism for enhanced immune response mediated by tumor epigenetic states.

## Results

### Baseline immune cell populations predict response and progression-free survival in sarcoma patients treated with immune checkpoint inhibitors

To study the influence of features linked to epigenetic states on antitumor immunity, we first characterized baseline immune infiltrates in tumor biopsies from 67 patients with a heterogeneous set of sarcomas (>10 subtypes) who were subsequently treated on ICI clinical trials (**Extended Data Table 1**). Twelve patients responded to ICI and 55 did not (CR/PR=12, SD=21, PD=34). Baseline samples were analyzed to identify tumor characteristics that could be informative prior to treatment and to eliminate confounding by varying ICI drugs and combinations used across trials. We employed an RNA sequencing (RNA-seq)-based method to quantify the abundance of different immune populations, MCP-counter^31^. To obtain robust clustering of samples based on their profile of immune infiltrates, we used a hierarchical clustering of principal components (HCPC) approach^32^, which integrates principal components (PCA) and hierarchical clustering. This HCPC revealed two highly distinct groups, which we deemed “immune-cold” and “immune-hot” (**Figure 1A, B; Extended Data Figure 1**). Except for cancer-associated fibroblasts, all cell types defined by MCP-counter were significantly associated with cluster partitioning, with T cells (p= 3.41 x 10^-10^) contributing the most, followed by cytotoxicity score (representative of cytotoxic lymphocytes) (p=2.01 x 10^-9^), and CD8^+^ T cells (p=6.89 x 10^-9^), NK cells (p=3.54 x 10^-8^), B cells (p=3.58 x 10^-8^), neutrophils (p=9.12 x 10^-8^), myeloid dendritic cells (p=1.17 x 10^-7^), macrophage/monocytes (p=1.34 x 10^-7^), and endothelial cells (p=7.88 x 10^-3^). The immune-hot cluster displayed, on average, greater abundance of all immune cell types in comparison to the overall mean, and conversely the immune-cold cluster displayed lower abundance of the same immune cell types (**Extended Data Table 2**).

**Figure 1.**
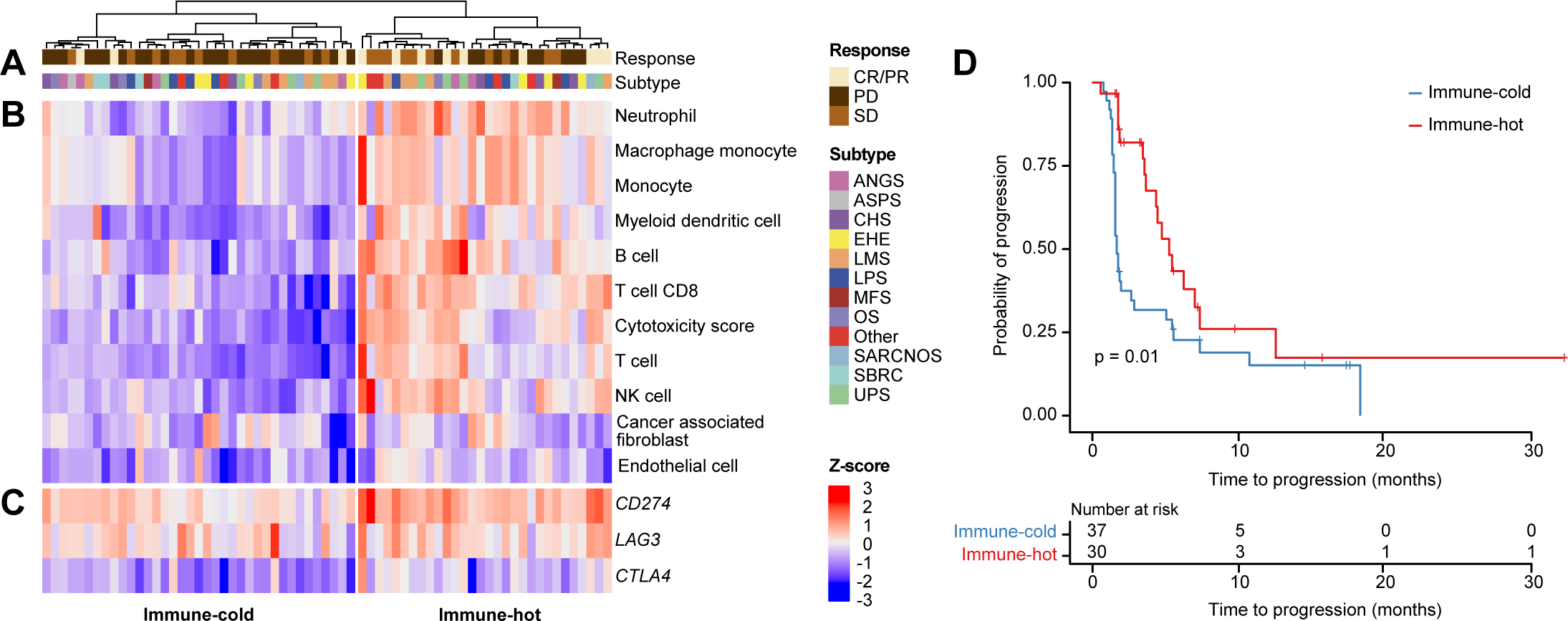
Clustering of immune cell fractions groups tumors into two distinct types. **A.** Color bars at the top of the heatmap label samples by response (SD, stable disease; PD, progressive disease; CR, complete response; PR, partial response), and histological subtype. Angiosarcoma, ANGS; alveolar soft part sarcoma, ASPS; chondrosarcoma, CHS; epithelioid hemangioendothelioma, EHE; leiomyosarcoma, LMS; liposarcoma, LPS; myxofibrosarcoma, MFS; osteosarcoma, OS; sarcoma not otherwise specified, SARCNOS; small blue round cell sarcoma, SBRC; undifferentiated pleomorphic sarcoma, UPS. **B.** Heatmap of immune and stromal cell fractions and cytotoxicity score determined by MCP-counter Z-scores. **C.** Immune checkpoint gene expression Z-scores. **D.** Kaplan-Meier plot representing progression-free survival (PFS) probability of immune-hot and -cold types. Tick marks indicate censoring. The P values on the Kaplan-Meier plots represent that output from cox proportional model that includes histology as a covariate.

Having assigned tumors to hot and cold immune groups, we next determined how these immune states correlated with clinical outcomes after the 67 patients in our cohort received ICI-based intervention in one of 3 clinical trials: pembrolizumab plus talmogene laherparepvec (NCT03069378)^33^, nivolumab plus bempegaldesleukin (NCT03282344)^2^, and pembrolizumab plus epacadostat (NCT03414229)^34^. There were no significant differences between the 3 ICI trials with respect to the number of responders and non-responders or immune-hot and -cold patients (**Extended Data Table 3**). We compared overall response rates (ORR) by RECIST version 1.1^35^ in immune-hot (ORR=30% [9/30]) vs. immune-cold (ORR=8.1% [3/37]) tumors. The ORR in the immune-hot group was significantly greater than in the immune-cold group (Fisher’s Exact Test, 95% CI 1.03-30.31, p=0.02). Furthermore, the immune hot samples were more prevalent than the immune cold samples in the complete response (CR) compared to progressive disease (PD) groups (Fisher’s Exact Test, 95% CI 0.02-0.73, p=0.01), while there was no significant difference in the CR versus stable disease (SD) and SD versus PD. The expression levels of immune checkpoint-related genes were consistent with the patterns observed in immune infiltrates, with elevated expression of *CD274* (PD-L1), *CTLA4* (two-sided t-test, p= 1.83 x 10^-6^ and p=1.18 x 10^-4^, respectively), and *LAG3* (two-sided t-test, p=0.12) in immune-hot tumors (**Figure 1C**).

To determine if the baseline immune type was prognostic for progression-free survival (PFS), we performed survival analysis including the histologic sarcoma subtype as a covariate (**Figure 1D, Extended Data Figure 2)**. Median PFS among patients with immune-cold tumors was 1.7 months vs. 3.65 months for immune-hot. Tumor classification as immune-hot contributed to improved PFS (HR=0.43, 95% CI 0.22-0.84, p=0.01) (**Extended Data Figure 2**). The histologic subtypes of leiomyosarcoma, myxofibrosarcoma, osteosarcoma, and small blue round cell tumors had a significant effect on PFS in our cohort.

### Hot and cold immune types are analogous to previously identified sarcoma immune classes

To determine how the two immune subtypes identified in this study relate to previously described sarcoma immune classes (SICs), which correlate with immune infiltrates and ICI response, we classified our samples according to those 5 SIC clusters (labeled A-E, **Extended Data Figure 3**)^19^. In total, 47% (14/30) of the immune-hot samples from our study fell into immune-hot SICs D and E. The remaining 53% (16/30) of immune-hot samples were assigned to immune-cold SIC B. In contrast, the immune-cold samples from our study were almost exclusively classified into immune-cold SICs A and B, with only two samples matching SIC C (**Extended Data Figure 3**). In summary, the two distinct immune clusters identified in this study between which we observe differences in PFS and ORR following ICI treatment are associated with SICs that are consistent with immune-high and immune-low states.

In addition to the validation of our clustering through comparison with independently developed classifications, we also reasoned that if the immune type clusters identified in our approach via deconvolution of bulk RNA sequencing accurately reflected immune cell populations, then the immune-hot cluster should contain more immune infiltrates than the immune-cold cluster, resulting in lower tumor content. Concordantly, the immune-hot type displayed significantly lower purity compared to the immune-cold type (two-sided t-test; t=-3.11, df=64, p=2.7 x 10^-3^) (**Extended Data Figure 4A**). To further confirm this relationship, we performed a permutation test randomly assigning samples to immune groups and comparing the difference between purity estimates between the two groups, which was repeated 10,000 times to produce a null distribution. The observed data displayed significantly greater differences in tumor purity estimates compared with the null distribution (p=3.1 x 10^-3^) (**Extended Data Figure 4B**). Lastly, tumor purity was inversely correlated with lymphoid and myeloid cell content (**Extended Data Figure 4C**), which is consistent with immune cell content contributing to the non-tumor cell fraction.

### TE and Ikaros (*IKZF1*) expression predict immune types in sarcoma

Although the activation of immune response through increased expression of transposable elements (TEs) and the involvement of epigenetic genes in the regulation of TE expression has been established in many cancers^23,24,36–39^, these processes have not been well studied in sarcoma. Our analysis of expression of 1,002 intergenic TEs across the two immune types shows heterogenous expression (**Extended Data Figure 5**). Thus, we next asked if expression of TEs and epigenetic regulators is predictive of tumor immune types in sarcoma. Lasso logistic regression models including expression of TEs (*R*^2^=0.29), epigenetic regulators (*R*^2^=0.19) had higher *R*^2^ values, indicating that the models including these features are a better fit for the prediction of immune type than a basic model that included only sarcoma subtypes and sequencing batch or models of TE and epigenetic regulator expression with randomized immune type sample labels (*R*^2^ TE shuffled=0.02, *R*^2^ epigenetic regulators shuffled=0.01) (**Figure 2A**). Furthermore, the selected models identified a small set of informative epigenetic regulator genes and TEs associated with the identified immune types from a large number of genes and TEs that were part of the model, i.e. signature features. Signature features with the highest contribution to the model included the MER57F (ERV1), MER45A (DNA transposon), Tigger17a (DNA transposon), MER61F (ERV1), LTR104_Mam (*Gypsy),* HERVL74.int (ERVL) TE subfamilies, expression of which was significantly greater in the immune-hot cluster (**Figure 2B** and **2C**). In addition, *IKZF1*, a chromatin-interacting transcription factor^40^ which regulates three-dimensional chromatin structure^41^, was the only epigenetic regulator of 532 genes tested as single genes to significantly contribute to the immune type prediction model and was associated with B cell infiltrates (**Figure 2B, 2C** and **Extended Data Figure 6A**).

**Figure 2.**
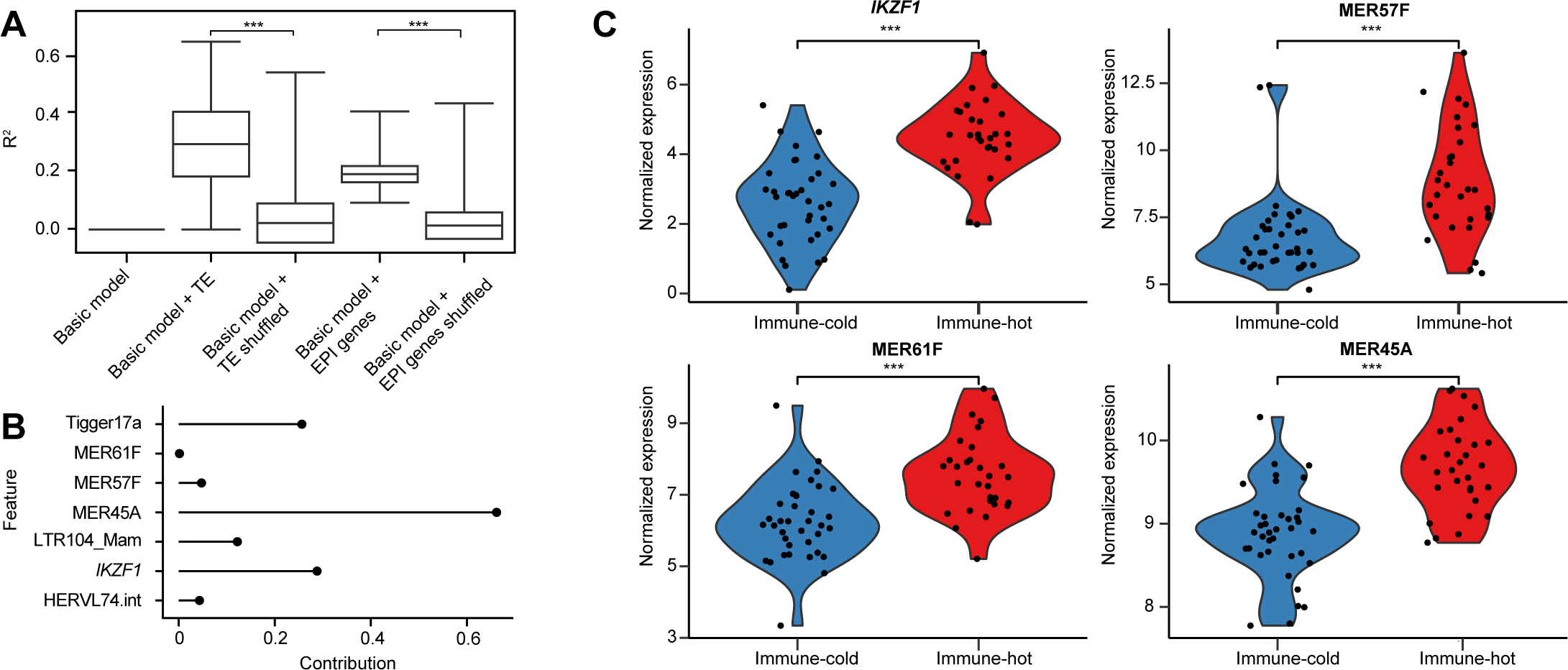
Transposable element and IKZF1 expression predict tumor immune groups. **A.** Comparison of lasso logistic regression model performances (R2) of the 5 tested models for prediction of immune type. P values determined by t-test; *** <2.2 x 10^-16^. **B.** Contribution of significant features from the TE and epigenetic models (models with the highest R2) represented as non-zero coefficients. The size and sign of contribution (coefficients) indicate the direction and strength of the feature’s effect on the outcome (immune cluster). **C.** Violin plots of normalized expression of transcripts identified as significant features in the regression model in immune-hot and -cold clusters. ***, p<0.001 as determined by one sided t-test.

We next fitted a logistic regression model using the signature features (i.e., *IKZF1* and TE score) to predict their effect on immune type. To calculate a TE score, we combined the expression values of the 6 signature feature TEs, for which expression of each of which was also positively correlated (**Extended Data Figure 6B**). After adjusting for sequencing batch and histology, we found that both TE score (p=2.2 x 10^-3^), and *IKZF1* expression (p=5.8 x 10^-3^), were significantly associated with immune type. This suggests that IKZF1 and TE affect immune type. We used a conditional independence approach (see Methods) to further investigate the potential causal relationships between *IKZF1*, TEs, and immune type. Our analysis revealed that: a) given IKZF expression, TE expression (TE score) is not conditionally independent of immune type (p=1.17 x 10^-5^), b) given TE expression, *IKZF1* expression is conditionally independent of immune type (p=0.14), and c) TEs and *IKZF1* do not have an independent impact on immune type (p=2.35 x 10^-6^). This analysis suggests that TEs play a significant role in determining immune type, and that they interact with IKZF1 in a complex way to modulate the immune response.

### High expression of TEs and *IKZF1* is associated with immune and inflammatory pathway signatures and progression free survival

We next determined whether *IKZF1* and TE expression correlated with activation of immune and inflammatory pathways using a partial Pearson correlation. Both *IKZF1* and TE score were positively correlated with multiple immune pathways, while pathways related to non-immune function were either significantly inversely correlated or not significantly correlated, suggesting a distinct relationship between TEs and *IKZF1* expression and immune activity in sarcomas (**Figure 3A**). Specifically, TE score and *IKFZ1* expression were significantly correlated with antiviral response pathways such as cGAS-STING (TE score, r^2^=0.64, p=7.90 x 10^-9^; *IKZF1*, r^2^=0.67, p=1.03 x 10^-9^), type I interferon (TE score, r^2^=0.38, p= 1.55 x 10^-3^, *IKZF1*, r^2^=0.32, p=7.89 x 10^-3^), and type II interferon (TE score, r^2^=0.68, p= 2.66 x 10^-10^, *IKZF1*, r^2^=0.55 p=1.28 x 10^-6^). Moreover, we observed positive correlations between TE score and *IKZF1* expression and the upregulation of antigen-processing machinery (TE score, r^2^=0.49, p=2.99 x 10^-5^, *IKZF1*, r^2^=0.27, p=2.44 x 10^-2^) as well as the CD8+ T cell effector pathway (TE score, r^2^=0.54, p=2.94 x 10^-6^, IKZF1, r^2^=0.45, p=1.17 x 10^-4^).

**Figure 3.**
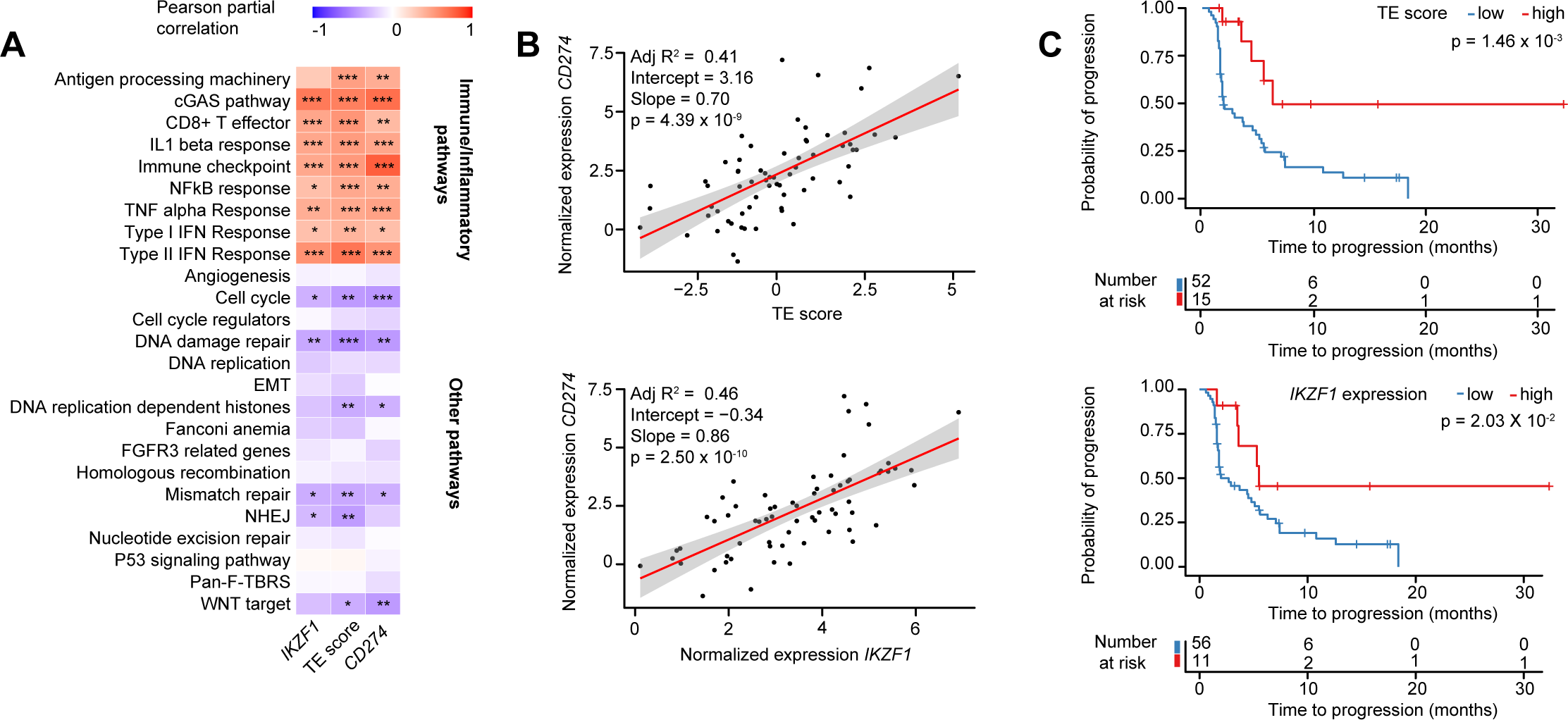
Immune pathway activation and progression-free survival following ICI treatment are associated with increased expression of multiple TE families and IKZF1. **A.** Heatmap of partial Pearson correlation including batch and histology as covariates. Scale from −1 (inverse correlation, blue) to 1 (positive correlation, red). Asterisks indicate Benjamini-Hochberg-corrected p-values: *p<0.05, ** p<0.01, *** p<0.001. **B.** Correlation between CD274 (PD-L1) gene expression and TE score, and CD274 and IKZF1 expression. **C.** Kaplan-Meier curves representing progression-free survival probability according to high vs. low TE scores and IKZF1 expression. The P values on the Kaplan-Meier plots represent that output from cox proportional model that includes histology as a covariate.

Because *CD274* (PD-L1) expression was significantly higher in the immune-hot group (**Figure 1C**), we also investigated the association between immune checkpoint-related genes and immune activity and found a significant positive correlation between *CD274* and immune and inflammatory pathways (CD8+ T cell effector, r^2^=0.36, adjusted p= 3.21 x 10^-3^, cGAS-STING, r^2^=0.71, p= 1.53 x 10^-11^; type II interferon, r^2^=0.54, p=2.51 x 10^-6^). TE score and *IKZF1* also positively correlated with *CD274* expression (p=4.40 x 10^-9^ and p= 2.5 x 10^-10^ respectively) (**Figure 3B**). We next tested whether the TE score and *IKZF1* expression were predictive of PFS. Both high TE score (p=1.65 x 10^-3^) and *IKZF1* expression (p=9.28 x 10^-3^) correlated with prolonged PFS (high TE 4.4 months vs. low TE 1.8 months; high *IKZF1* 5.3 months vs. low *IKZF1* 1.8 months) (**Figure 3C**). The ORR based on *IKZF1* expression was 54.5% (6/11) in the high-expressing group and 10.7% (6/56) in the low-expressing group (p=2.72 x 10^-3^; Fisher’s exact test). ORR in the TE-high group was 40% (6/15) and 11.53% (6/52) in the TE-low group (p=0.13; Fisher’s exact test). Taken together, these findings suggest that both *IKZF1* expression and TE score, which were identified in model that considered subtypes as a variable, could be explored as predictive biomarkers for ICI outcomes.

### TE and *IKZF1* expression associate with immune infiltrate and inflammatory pathway activation in a separate validation cohort of sarcoma patients

To assess the replicability of our findings, we applied our analysis to gene expression data from 190 sarcoma samples from the TCGA^3^. This group was chosen as it includes 5 sarcoma subtypes, DDLPS (n=49), MFS (n=17), LMS (n=80; 53 STLMS; 27 ULMS), and UPS (n=44), which were prevalent in our original cohort. The immune signatures in the validation cohort segregated into two distinct clusters marked by high (immune-hot) and low (immune-cold) immune infiltrates and expression of immune checkpoints (**Figure 4A-C, Extended Data Figure 7**). The immune-hot cluster was associated with improved overall survival (p=1.09 x 10^-2^), at a median of 37.5 months vs. 25.5 months for the immune-cold cluster (**Figure 4D**).

**Figure 4.**
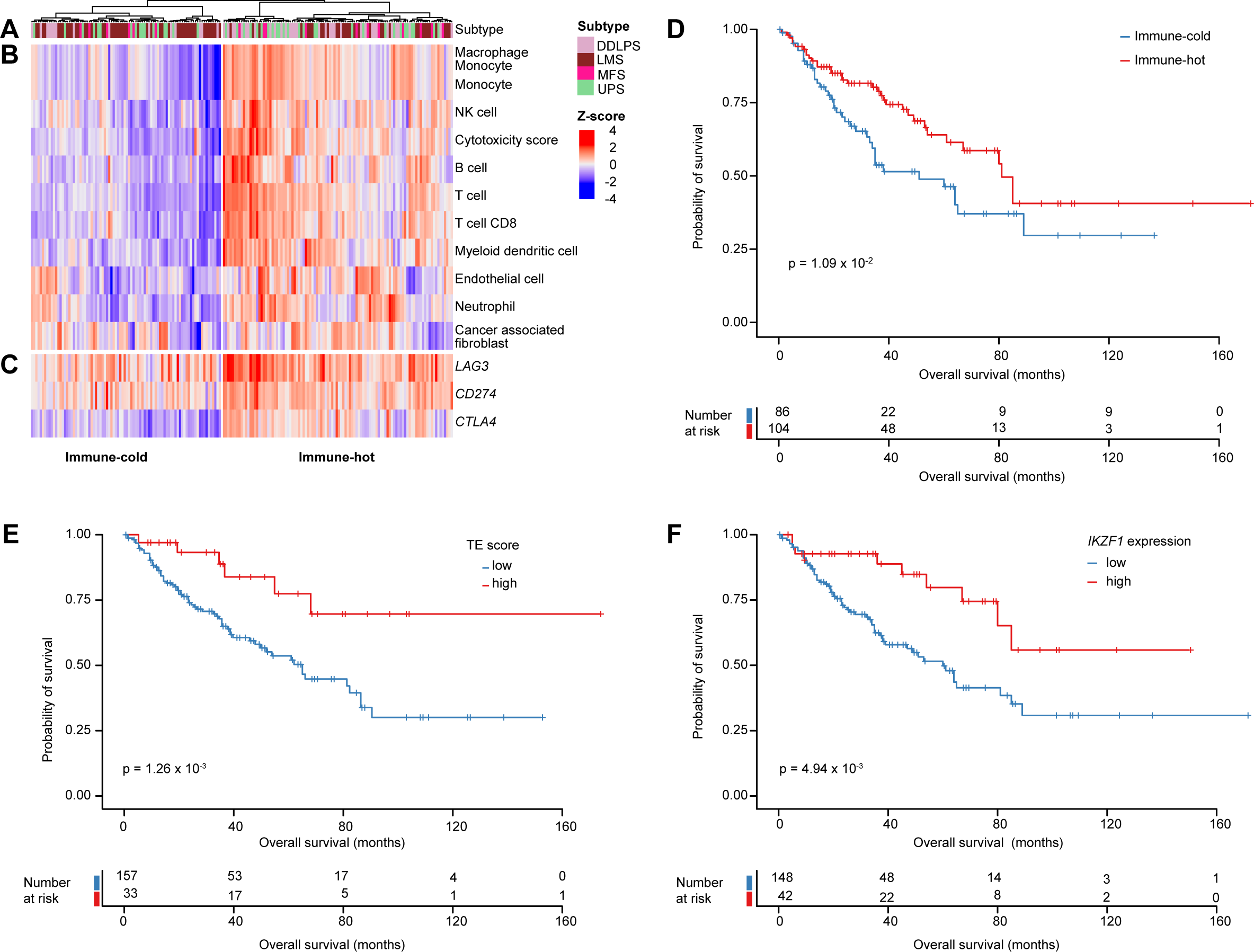
TE score and IKZF1 expression associate with improved survival in a validation cohort. A. Clustering of samples into immune-hot and immune-cold types. Color bar at top labels samples by histological subtype. DDLPS, dedifferentiated liposarcoma; LMS, leiomyosarcoma; MFS, myxofibrosarcoma; UPS, undifferentiated pleomorphic sarcoma. **B.** Heatmap of immune and stromal cell fractions and cytotoxicity score determined by MCP-counter Z-scores. **C.** Immune checkpoint gene expression Z-scores. **D.** Kaplan-Meier plot of overall survival probability of patients with immune-hot and -cold type tumors. **E-F.** Kaplan-Meier curves representing overall survival probability of high (red) and low (blue) **E.** TE scores and **F.** high (red) and (low) IKZF1 expression. The P values on the Kaplan-Meier plots represent that output from cox proportional model that includes histology and tumor size as covariates.

As in the original cohort, expression of TEs and epigenetic regulators predicted immune type (**Extended Data Figure 8A**) and specific TEs and *IKZF1* were identified as signature features that positively correlated with immune-hot classification (**Extended Data Figure 8B, C**). Furthermore, expression of *IKZF1* and TEs (again defined as a composite TE score) correlated with that of immune pathways including type I and II interferon (p<0.001), antigen-processing machinery (p<0.001), and immune checkpoint genes (p<0.001) including *CD274*, but not non-immune pathways (**Extended Data Figure 9A, B**). Overall survival was greater for patients whose tumor had a high TE score (p=1.26 x 10^-3^) or *IKZF1* (p=4.94 x 10^-3^) expression (**Figure 4E, F**).

## Discussion

To address the pressing need to identify predictive biomarkers of response to ICI-based therapy in sarcomas, we identified the minimal number of immune clusters that represent immune-hot and -cold sarcomas and showed that the former is associated with higher ORR and longer PFS following ICI treatment independent of subtype. This finding corroborates prior studies that have shown a correlation between high baseline immune infiltrates and response to immune therapy^19^. Importantly, our work demonstrates that these findings apply in a cohort with a broad spectrum of sarcoma subtypes and in the setting of 3 combination ICI trials with diverse mechanisms. Moreover, our analysis shows that a binary classification of tumors is sufficient to correlate with clinical outcomes, indicating that immune clustering can be simplified compared to previous approaches that involved more groups^19^. Such a simplified system could be helpful in smaller studies with limited numbers of cases.

To identify specific tumor-intrinsic features that contribute to differences in immune states, we focused on epigenetic regulation. Epigenetic mechanisms are known to suppress antitumor immune responses and targeting epigenetic pathways has emerged as a promising therapeutic strategy^21,30,42^. Specifically, we examined the expression of epigenetic regulators and TEs, the latter of which are normally epigenetically silenced (e.g., via establishment of heterochromatin) and can stimulate innate immune responses when de-repressed^27–29,43^. We observed increased expression of TEs in immune-hot tumors. This is consistent with the ability of TEs to activate dsRNA-sensing pathways, as has been observed in the setting of genetic lesions in epigenetic regulators or pharmacologic treatment that lead to their de-repression^23,24,30,44^. Our observation of upregulated antiviral immune responses (including cGAS and type I interferon signaling) and antigen-presenting pathways is consistent with this mechanism. Further investigation is needed to determine whether the presentation of TE-derived neoantigens via MHC-I, as observed in the loss of epigenetic TE silencing, could also contribute to the immune-hot state^27,38^.

In addition to TEs, our analysis revealed that expression of *IKZF1* was significantly greater in immune-hot tumors and associated with PD-L1 expression and B cell infiltrates, which was validated in a separate cohort of 190 sarcoma samples from the TCGA. Notably, greater infiltration of B lineage immune cells associates with overall survival in soft tissue sarcomas^19^. Although Ikaros, the *IKZF1* gene product, is primarily studied as a transcription factor in hematologic lineages^40^, it was included in our list of epigenetic regulators given the inherent interaction of transcription factors and chromatin. Recent reports also suggest an important role for Ikaros in regulating higher order chromatin structure^41^. Notably, previous studies have determined that if *IKZF1* is expressed in tumor cells and not only in immune populations, it contributes to upregulation of immune infiltration and enhances the efficacy of anti-PD-1 and anti-CTLA-4 immunotherapies in murine models^45^. Our findings raise the possibility of a similar effect in sarcomas.

It is unclear whether TE de-repression and *IKZF1* expression are directly linked mechanistically. One possibility is that IKZF1 regulates TE expression, as several TE families contain an IKZF1-binding motif^46^. It is also possible that the mechanism for loss of epigenetic silencing at TEs creates a permissive chromatin state for *IKZF1* binding that allows for activation of nearby genes involved in innate immune activation. Alternatively, de-repression of TEs, which can act as cis-regulatory elements, could promote *IKZF1* expression. In our study, conditional independence analysis supports a model in which IKZF1 regulates TE expression, which in turn determines immune types. However, functional studies are needed to test this hypothesis.

There are several limitations of this study including the relatively small sample size (n=67), the heterogeneity in sarcoma subtypes, and that patients were included from 3 trials of ICI-based regimens with different mechanisms. However, while this heterogeneity may have decreased our ability to identify signals related to specific epigenetic genes or TE families, we were reassuringly able to classify tumors into immune classes that were predictive of clinical outcomes and confirm prior classification systems. Furthermore, our key findings were confirmed in a larger validation cohort (n=190), which included common sarcoma subtypes also represented in the original sample set. However, the validation cohort differed in that samples were from patients who had not received systemic therapy, it was composed of nearly all primary tumors, and outcome was overall survival and not PFS or response following ICI-based treatment. Another caveat of the study is that we selected polyadenylated transcripts for RNA sequencing, which would limit detection of theoretically transcribed but non-polyadenylated TEs. We were also potentially limited by considering epigenetic genes as independent, when many encode proteins that form complexes or functional pathways.

Increasing the effectiveness of immunotherapies and identifying predictors of ICI response would both represent important advances in sarcoma. Our work presents several possibilities for achieving these goals using data from pretreatment biopsies. We confirm earlier studies showing that pretreatment immune status can predict ICI outcomes and propose *IKZF1* expression and TE score as potential predictive biomarkers for ICI response, both of which require validation. In addition, this work reveals potential avenues to enhance ICI response through stimulation of immune responsiveness of baseline immune-cold tumors to convert them into an immune-hot phenotype. Based on this work, promoting the de-repression of TEs by pharmacologic targeting of epigenetic regulators could be explored in preclinical models.

## Materials and Methods

Clinical data were collected and DNA and RNA sequencing of pre-treatment biopsy samples was performed under Institutional Review Board oversight of 3 clinical trials performed at the Memorial Sloan Kettering Cancer Center. These include pembrolizumab plus talmogene laherparepvec (NCT03069378)^33^, nivolumab plus bempegaldesleukin (NCT03282344)^2^, and pembrolizumab plus epacadostat (NCT03414229)^34^. Details regarding each study’s design, safety oversight, and interventions can be found in referenced publications for each study.

### Samples

A total of 67 baseline samples from twelve sarcoma subtypes (angiosarcoma (ANGS) =4, alveolar soft part sarcoma (ASPS)=1, chondrosarcoma (CHS)=6, epithelioid hemangioendothelioma (EHE)=8, leiomyosarcoma (LMS)=11, liposarcoma (LPS)=8, myxofibrosarcoma (MFS)=2, osteosarcoma (OS)=4, Other=7, sarcoma not otherwise specified (SARCNOS)=2, small blue round cell sarcoma (SBRC)=4, and undifferentiated pleomorphic sarcoma (UPS)=8, representing twelve responders and 55 non-responders (CR/PR=12, SD=21, PD=34) were transcriptionally profiled (**Extended Data Table 1**).

### RNA sequencing, and quantification of TEs and genes

After quantification of RNA using RiboGreen and quality control using the Agilent BioAnalyzer, 469-500 ng of total RNA with RNA integrity values ranging from 6.8–10 underwent polyA selection and TruSeq library preparation following the instructions provided by Illumina (TruSeq Stranded mRNA LT Kit, catalog #RS-122-2102), with 8 cycles of PCR. The resulting samples were barcoded and run on a HiSeq 4000 at 100 paired-end reads, using the HiSeq 3000/4000 SBS Kit (Illumina), generating an average of 41 million paired reads per sample. Ribosomal reads represented 0.9–5.9% of the total reads generated and the percent of mRNA bases averaged 64%.

The obtained FASTQ files were processed using the REdiscoverTE^38^ workflow, which allowed for quantification based on transcript levels. Gene transcripts were aggregated to obtain individual gene quantification. Read counts for each individual transposable element (TE) were then gathered to the level of TE subfamily, family, and class, as defined by the human Repeatmasker Hg38. TE expression was further divided into inter- and intragenic regions as defined by Gencode GTF/GFF and implemented in REdiscoverTE. Downstream analysis considered only intergenic expression of 1002 out of a total of 1052 TE subfamilies that were expressed. Gene-based normalization factors were calculated using the ‘RLE’ algorithm in edgeR^47^, as determined by REdiscoverTE. The data was further variance-stabilized using the voom function from edgeR.

### RNAseq deconvolution and generation of immune clusters

We quantified immune cell populations from variance-stabilized RNAseq data using the immunedeconv R package and its deconvolute function, along with the MCPcounter option 3.6.3^31^. Batch effects due to sequencing run were removed using removeBatchEffect() function from the limma R package^48^. To reduce the dimensionality of the immune cell proportion data, we first performed a principal component analysis, followed by hierarchical clustering on principal components (HCPC) using the FactoMineR package ^49^. Cluster types were visualized using the factoextra R package (https://CRAN.R-project.org/package=factoextra).

Heatmaps of expression in each cluster were generated based on the scaled (Z-scores) immune cell proportions. Z-scores were calculated using the formula z = (x-μ)/σ, where x is the raw cell fraction, μ is the mean of all samples, and σ is the standard deviation for all samples.

To obtain the cellularity enrichment scores for 64 cell types, from which lymphoid and myeloid cell type proportions can be derived, we used the xCellAnalysis function in the xCell R package (https://github.com/dviraran/xCell). Total lymphoid content was calculated as the sum of 21 lymphoid cell scores, including CD8 + T cells, NK cells, CD4 + naive T cells, B cells, CD4+ T cells, CD8+ Tem, Tregs, plasma cells, CD4 + Tcm, CD4+ Tem, memory B cells, CD8+ Tcm, naive B-cells, CD4+ memory T cells, pro B cells, class-switched memory B cells, Th2 cells, Th1 cells, CD8+ naive T cells, NKT, and Tgd cells. Total myeloid content was expressed as the sum of 13 cell scores, including monocytes, macrophages, dendritic cells (including activated, conventional, interstitial, and plasmacytoid), neutrophils, eosinophils, M1 macrophages, M2 macrophages, basophils, and mast cells.

To test if immune type predicts survival in sarcoma, we performed Cox regression analysis that included histology to control for subtype-specific differences in outcomes. We compared survival between groups using the Kaplan-Meier survival curve and the Cox proportional-hazards regression mode. Differences were considered significant if the p-value was less than 0.05 for the tested group.

### Exome sequencing and purity estimation

Viably frozen cells were thawed and pelleted and incubated for at least 30 min in 360 μL Buffer ATL + 40 μL proteinase K at 55°C. DNA was isolated using the DNeasy Blood & Tissue Kit (QIAGEN catalog #69504) according to the manufacturer’s protocol with 1 h of incubation at 55°C for digestion. DNA was eluted in 0.5X Buffer AE.

After PicoGreen quantification and quality control by Agilent BioAnalyzer, 100-250 ng of DNA was used to prepare libraries using the KAPA Hyper Prep Kit (Kapa Biosystems KK8504) with 8 cycles of PCR. After sample barcoding, 100-500 ng of library DNA was captured by hybridization using the xGen Exome Research Panel v1.0 (IDT) according to the manufacturer’s protocol. Post-capture libraries were amplified using 8 PCR cycles. Samples were run on a HiSeq 4000 at 100 paired-end reads using the HiSeq 3000/4000 SBS Kit (Illumina). Normal and tumor samples were covered to an average of 102X and 219X, respectively.

FASTQ files were aligned and processed using the in-house workflow Tempo (https://github.com/mskcc/tempo)^50^. Briefly, reads were aligned using Burroughs-Wheeler Aligner (BWA)-MEM ^51^ to the GRCh37 reference genome and base recalibration was performed using Genome Analysis Toolkit (GATK) best practices. Somatic genome variants were called using the union of Mutect2 and Strelka2. Variants were then filtered based on the following criteria: tumor read depth of 20, variant allele frequency <0.5 x the tumor alternate read count of 3, and normal read depth of 10. In addition, repeated regions from RepeatMasker^52^ and variants that appear at allele frequencies >0.01 in GNOMAD^53^ were filtered out. Somatic copy number alterations were analyzed using FACETS (Fraction and Allele-Specific Copy Number Estimates from Tumor Sequencing) v0.5.14^54^. Each tumor and matched normal pair was processed in two steps: a first run for ploidy and purity estimation followed by a second run for detection of focal events. Each fit was reviewed manually to minimize false positives and to estimate the quality of the fit. Purity estimates from facets were used in the subsequent analysis.

### Lasso association between immune types and genomic features

To identify genomic features that significantly differed between the two immune types, we used lasso logistic regression via penalized maximum likelihood using the R package *glmnet*^55^. To account for potential variations due to sequencing batch or subtypes, we included these parameters in the basic model. Other models were further built with either normalized expression of 1002 intergenic TEs, shuffled TE expression, normalized expression of epigenetic modulators (532 genes), or shuffled epigenetic modulator expression.

1. Basic model: Immune types∼ batch + sarcoma subtypes
2. Basic model + TE: Immune types∼ batch + sarcoma subtypes + 1002 TEs
3. Basic model + TE shuffled: Immune types∼ batch + sarcoma subtypes + 1002 TEs shuffled
4. Basic model + epigenetic genes: Immune types∼ batch + sarcoma subtypes + 532 epigenetic genes
5. Basic model + epigenetic genes shuffled: Immune types∼ batch + sarcoma subtypes + 532 epigenetic genes shuffled

TEs in the models represent intergenic TEs, and shuffled TE or epigenetic genes data represents randomly assigned TE or epigenetic genes expression to the samples.

Tenfold cross-validation was performed for each regression, and lasso coefficients at one standard error of the minimum mean cross-validation errors (lambda 1se) were used. Each lasso fit returned a small number of predictors, i.e. variables with non-zero coefficients, matching genomic features with significant contributions to difference between the two immune types. *R*^2^ values for each model were calculated from the fraction of deviance explained and averaged across the 10 rounds of cross-validation. *R*^2^ values were then used to determine the model with the best performance. To identify notable features associated with immune type, we extracted non-zero coefficients of the final best models.

To further test the relationship between significant TE and epigenetic features determined by the glmnet model, we used logistic glm regression in which immune type represented a dependent variable, while TE score and *IKZF1* expression represented independent variables. The model was corrected for batch and histology covariates. The TE score was calculated by generating a z-score for the expression of 8 TEs found to be significant in the glmnet analysis and that exhibited positive correlation with each other (**Extended Data Figure 6B**). Z-score was generated using the gsva function of the GSVA package^56^.

### Logistic regression and conditional independence test

To further confirm the relationship between selected TEs and *IKZF1* expression with respect to immune cluster, we performed a logistic regression test. TE score and *IKZF1* were used as independent variables to assess their association with immune type. Bach and histology were used as covariates in the model.

Conditional independence (mutual information) tests to identify causal relationships between TEs, IKZF1, and immune-hot/-cold phenotype were performed using the bnlearn package in R (https://www.bnlearn.com/). Three hypotheses were tested:

a. TE score -> *IKZF1*-> Immune type: Immune type is conditionally independent of TE given *IKZF1*; TEs regulate
b. *IKZF1* and do not directly regulate immune type. *IKZF1*-> TE score -> Immune type: Immune type is conditionally independent of *IKZF1* given TE; *IKZF1* regulates TEs and does not directly regulate immune type.
c. *IKZF1*-> Immune type <-TE score: *IKZF1* and TEs are conditionally independent of immune type.

### Gene signature calculations

Genes for immune/inflammatory and other signatures used to determine the correlation of significant features found to be predictive of immune type were defined as previously described in literature and summarized in Kong et al.^38^ (except the cGAS pathway, which was downloaded from KEGG https://www.gseamsigdb.org/gsea/msigdb/cards/). The ssGSEA algorithm was used to comprehensively assess gene signature expression of each^57^. The correlation between gene signatures and normalized expression of significant features was assessed by partial Pearson correlation analysis with batch and histology as covariates. P values were corrected using the Benjamini-Hochberg correction.

### Comparison with previously reported immune classes

To compare our immune clusters with formerly derived 5 sarcoma immune classes (SIC) previously defined by Petitprez et al.,^19^ we obtained centroid infiltration scores for each of 4 cell types (i.e. T cells, cytotoxic scores, B lineage, endothelial cells) of the 5 clusters derived from MCP-counter analysis from the authors of the paper. We then calculated Euclidian distance (distance = √Σ(A_i_-B_i_)^2^) between centroids of four cell types (i.e. T cells, cytotoxic scores, B lineage, endothelial cells) from each SIC (i.e. A,B,C,D,E) and the Z-score scaled MCP-counter proportions from the same four cell types in our data. Each sample was assigned to SIC type based on the lowest Euclidian distance with the 4 centroid infiltration scores for each SIC. Z-score-scaled immune cell proportions were then plotted using the Complex heatmap package in R, and the comparison with our Immune hot and cold clusters was performed.

### TCGA data analysis

RNA sequencing data and phenotypic information were obtained from dbGaP for 190 TCGA samples from 5 sarcoma subtypes, including DDLPS (n=49), MFS (n=17), LMS (n=80; 53 STLMS +27 ULMS), and UPS (n=44). The REdiscoverTE pipeline was used to quantify gene and TE expression. Batch effect information was downloaded from the TCGA Batch Effects Viewer (https://bioinformatics.mdanderson.org/public-software/tcga-batch-effects/) and considered in the subsequent data analysis. RNA sequencing was deconvoluted, immune clusters identified, and lasso associations between immune types and genomic features and overall survival were analyzed as described above. For the Kaplan-Meier analysis of this dataset our Cox regression analysis included histology and tumor size. The latter was included since the TCGA dataset comprises nearly all primary cases in which tumor size can be an more important prognostic factor.

## Data Availability

All RNA sequencing data, where informed consent has been obtained from the patient, is publicly available via dbGaP (accession number: phs003284). Three samples are not publicly available due to lack of consent for their release. All exome recapture sequencing data will be available via dbGaP under accession number phs001783 by the time of publication.

## Code Availability

Custom code used for analysis is publicly available here: https://github.com/BradicM/Sarcoma_TE_paper_analysis

## Acknowledgements

This work was supported by Merck, Amgen, NEKTAR, Incyte, Bristol Myers Squibb, Cycle for Survival, and Witherwax Fund. BAN received support from the NCI K08CA245212, the Connective Tissue Oncology Society Basic Science Research Award (with WDT), and the Damon Runyon Clinical Investigator Award. Additional support was provided by the Memorial Sloan Kettering Cancer Center Support Grant (P30 CA008748) and Hillman Cancer Center Support Grant (P30 CA047904) provided additional support. We acknowledge the use of the Integrated Genomics Operation Core, funded by the NCI Cancer Center Support Grant (CCSG, P30 CA08748), and the Marie-Josée and Henry R. Kravis Center for Molecular Oncology.

---

**Extended Data Figure 1.**
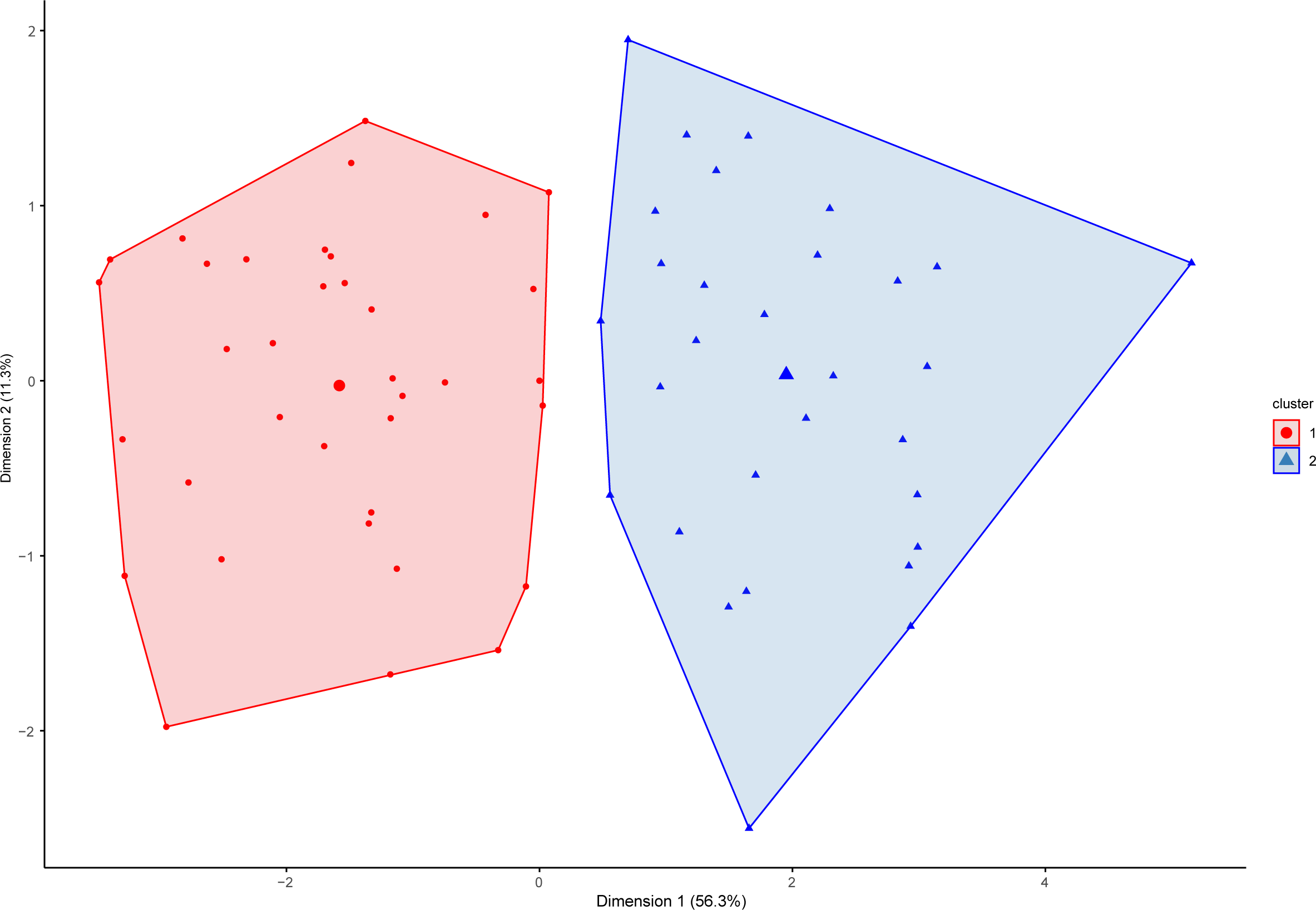
Determination of immune clusters from MCP-counter-based immune-deconvoluted cell proportions. Factor map representing two clusters based on hierarchical clustering of principal components. Each dot represents an individual patient sample.

**Extended Data Figure 2.**
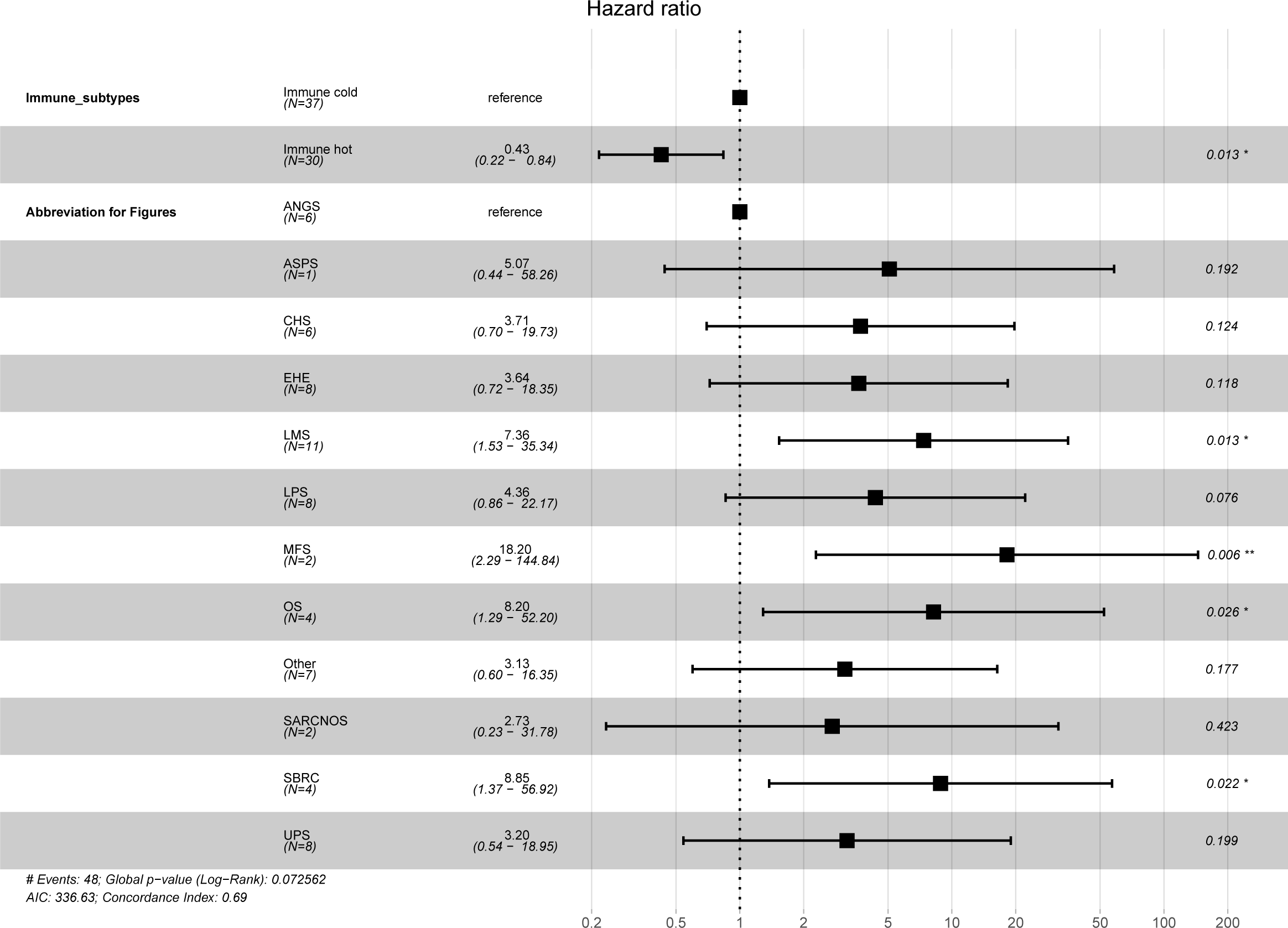
Forest plots showing multivariable Cox regression analysis of contribution of immune cluster, histology, and clinical protocol to risk of progression. p-values calculated using Cox proportional hazards analysis (*, p ≤ 0.05).

**Extended Data Figure 3.**
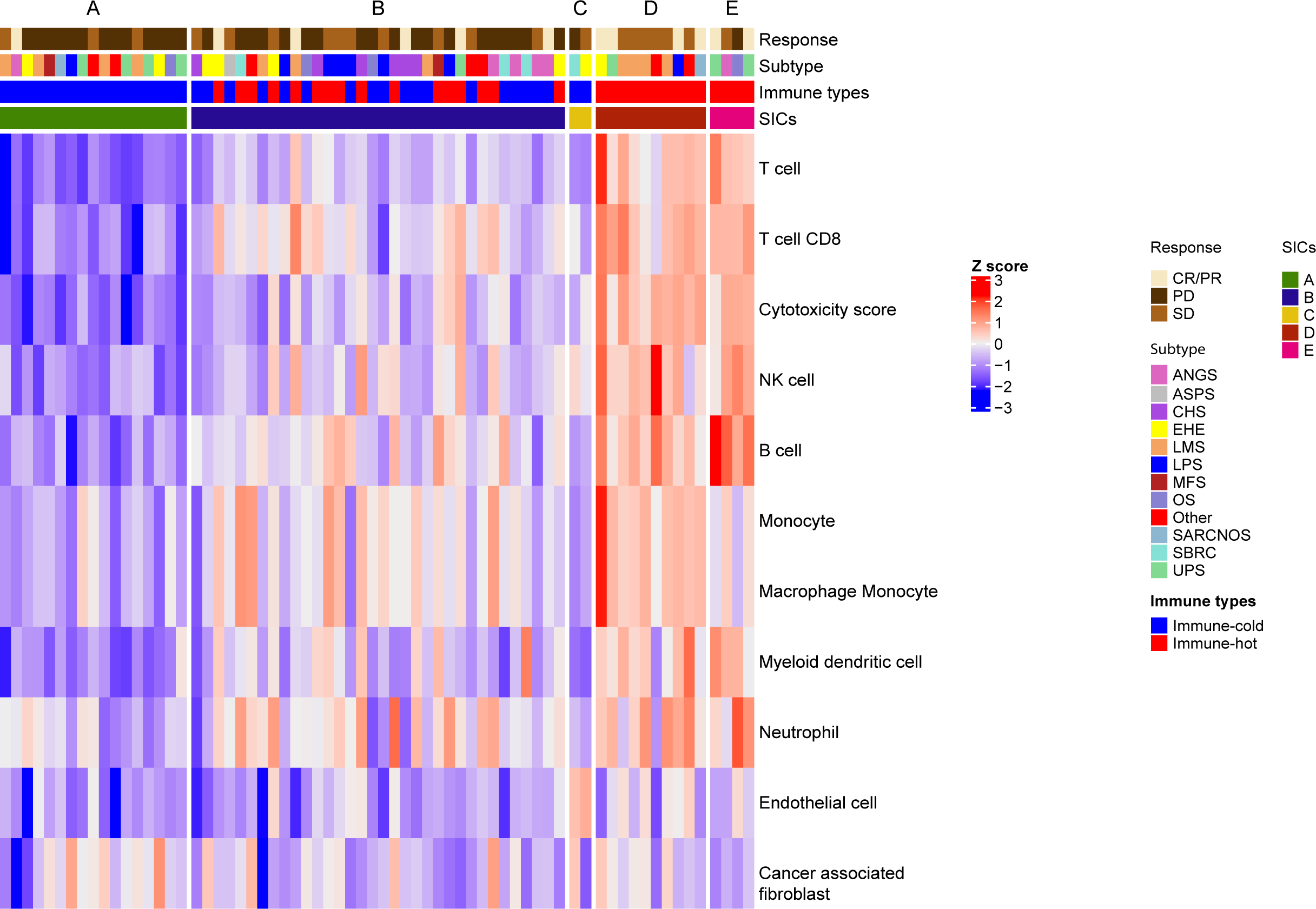
Hot and cold immune types are related to previously identified sarcoma immune classes. SIC types of each sample in study cohort as determined by the Euclidian distance between centroids of the proportions of 4 cell types (i.e. T cells, cytotoxic scores, B lineage, endothelial cells) between each SIC (i.e., A, B, C, D, E)19 and proportions in each sample. Color bars at top of heatmap label the samples by response, histological subtype, SIC, and immune type. Heatmap indicates immune cell fraction determined by MCP-counter Z-score.

**Extended Data Figure 4.**
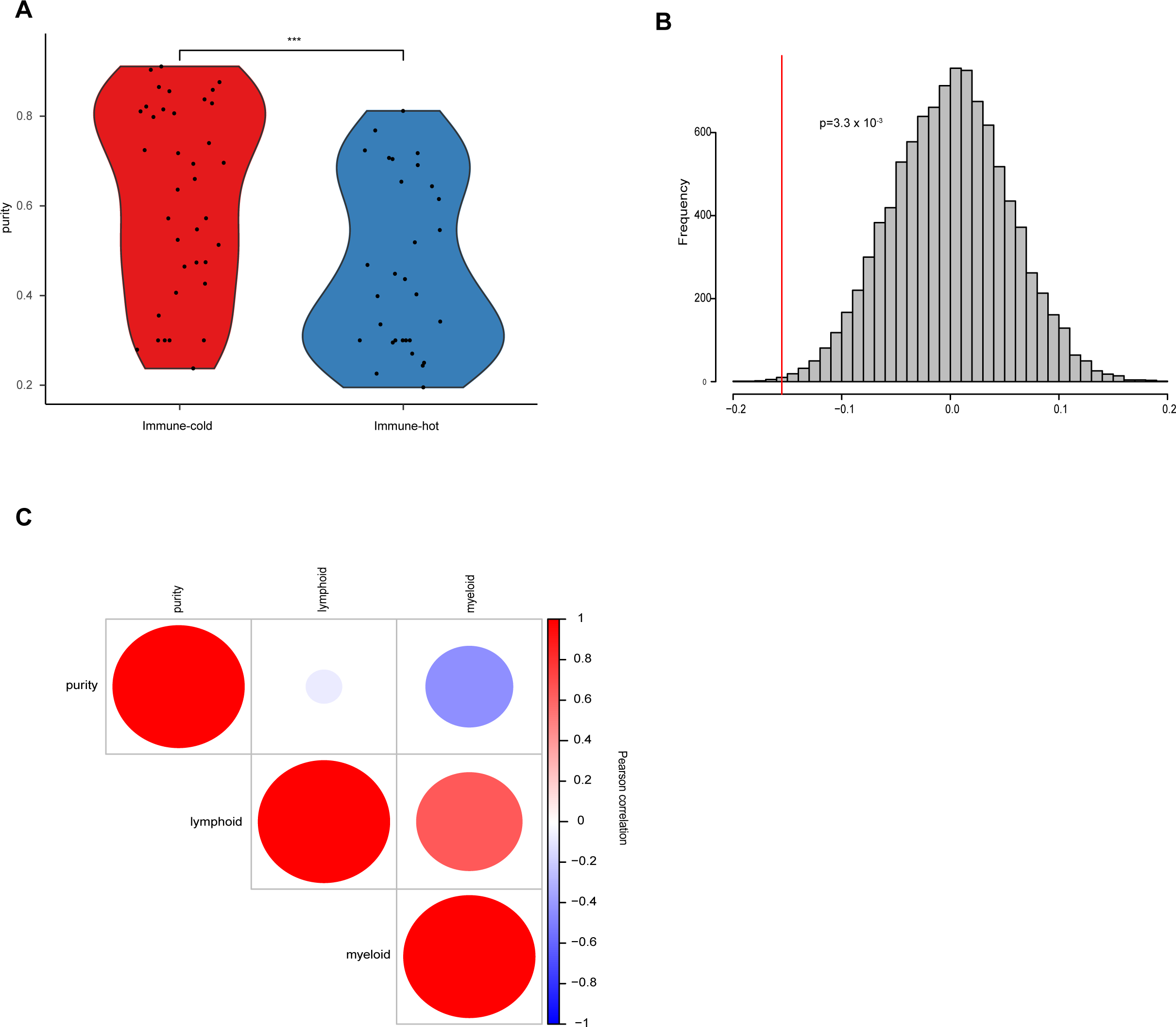
Immune infiltrates are inversely related to sample purity. **A.** Violin plot comparing tumor purity between the two immune types. **B.** Distribution of 10,000 permutations of tumor purity shuffling between “immune-cold” and “immune-hot. The histogram shows the simulated absolute permuted differences in means. The vertical red line represents the observed value for the original two samples (immune-cold and immune-hot). **C.** Correlation between purity, lymphoid content, and myeloid cell content. Scale from −1 (inverse correlation, blue), to 1 (positive correlation, red). Areas of circles represent the absolute value of corresponding correlation coefficients.

**Extended Data Figure 5.**
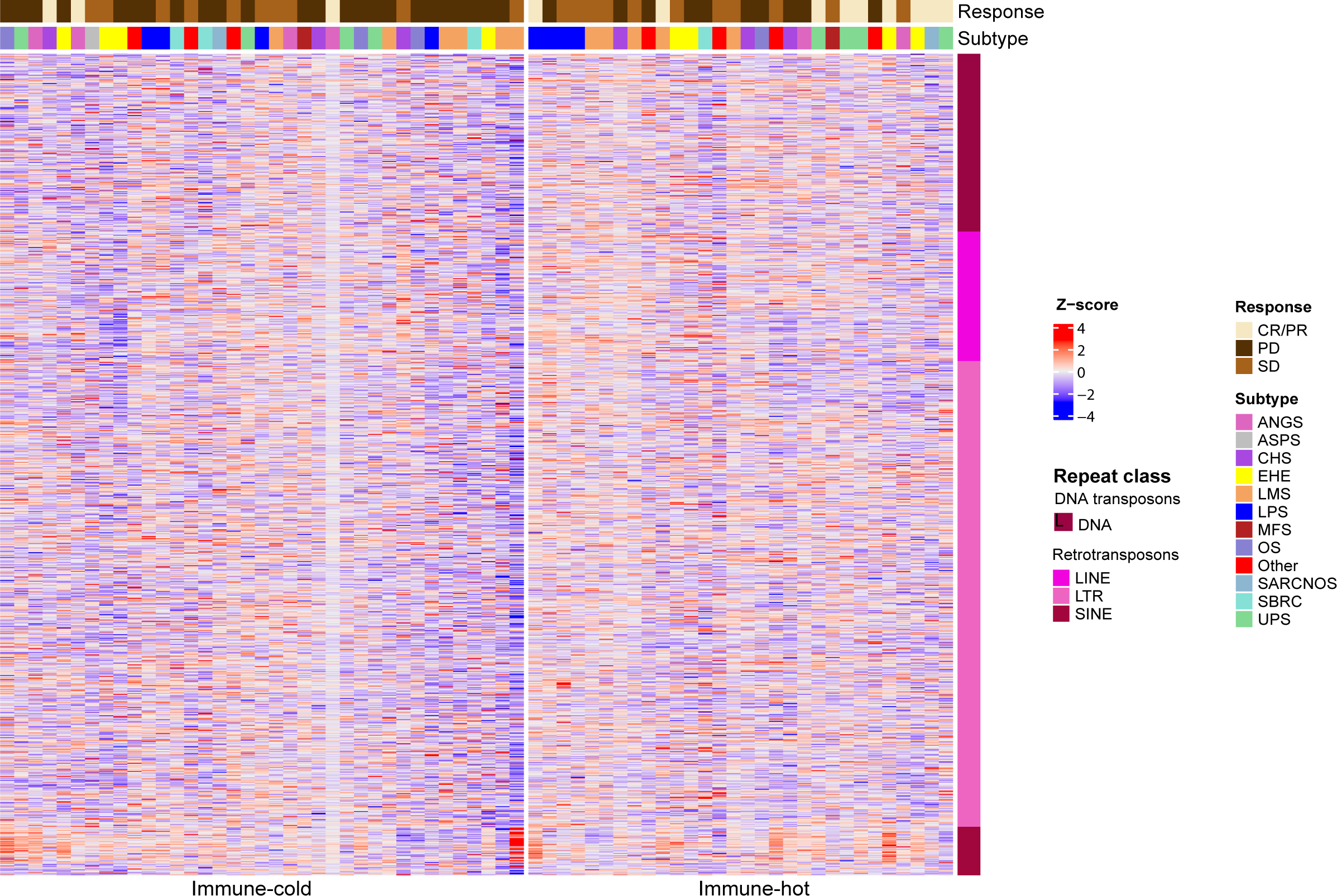
TE expression is heterogenous across sarcoma samples. Expression of all 1002 intergenic TEs expressed in the studied samples. Color bars at top of heatmap label the samples by response and histological subtype. Color bar at right labels repeat classes; LINE-Long interspersed nuclear elements, LTR-long terminal repeats, SINE-short interspersed nuclear elements. TE expression represented as Z-score; batch effect was removed.

**Extended Data Figure 6.**
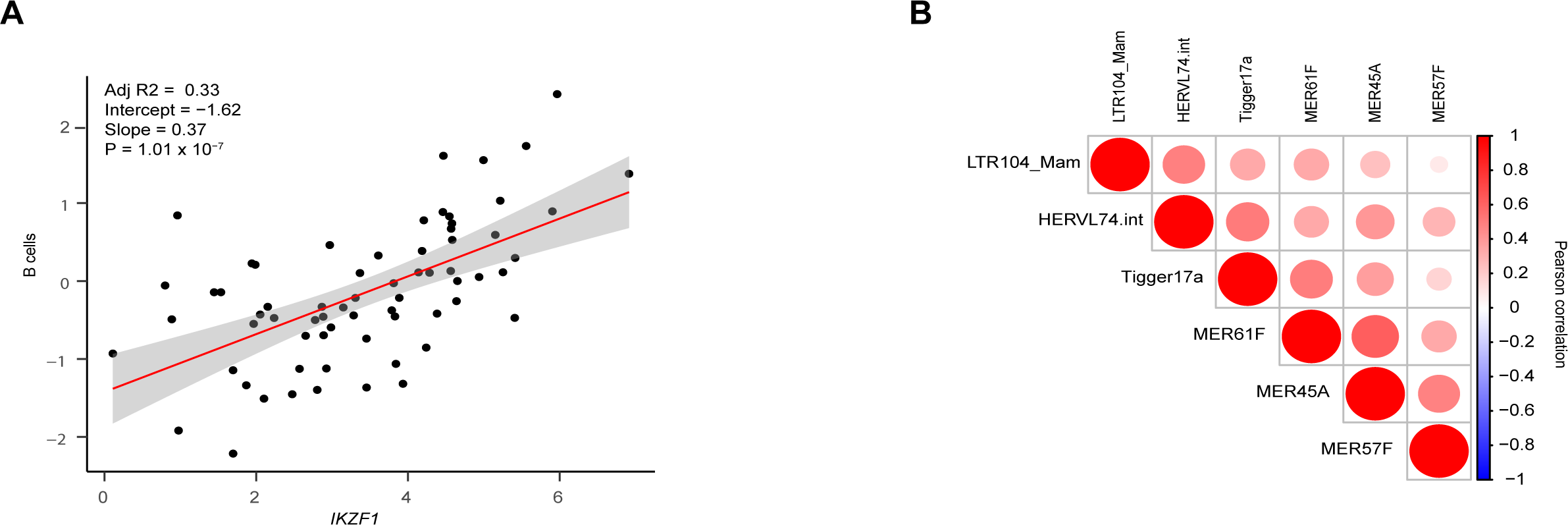
Correlation between IKZF1 expression and B cell infiltrates and between eight significant TEs detected in our model. **A.** Correlation between IKZF1 expression and B cell infiltrates. **B.** Pearson correlation among expression of 6 TEs. Scale from −1 (inverse correlation, blue) to 1 (positive correlation, red). Areas of circles represent the absolute value of corresponding correlation coefficients.

**Extended Data Figure 7.**
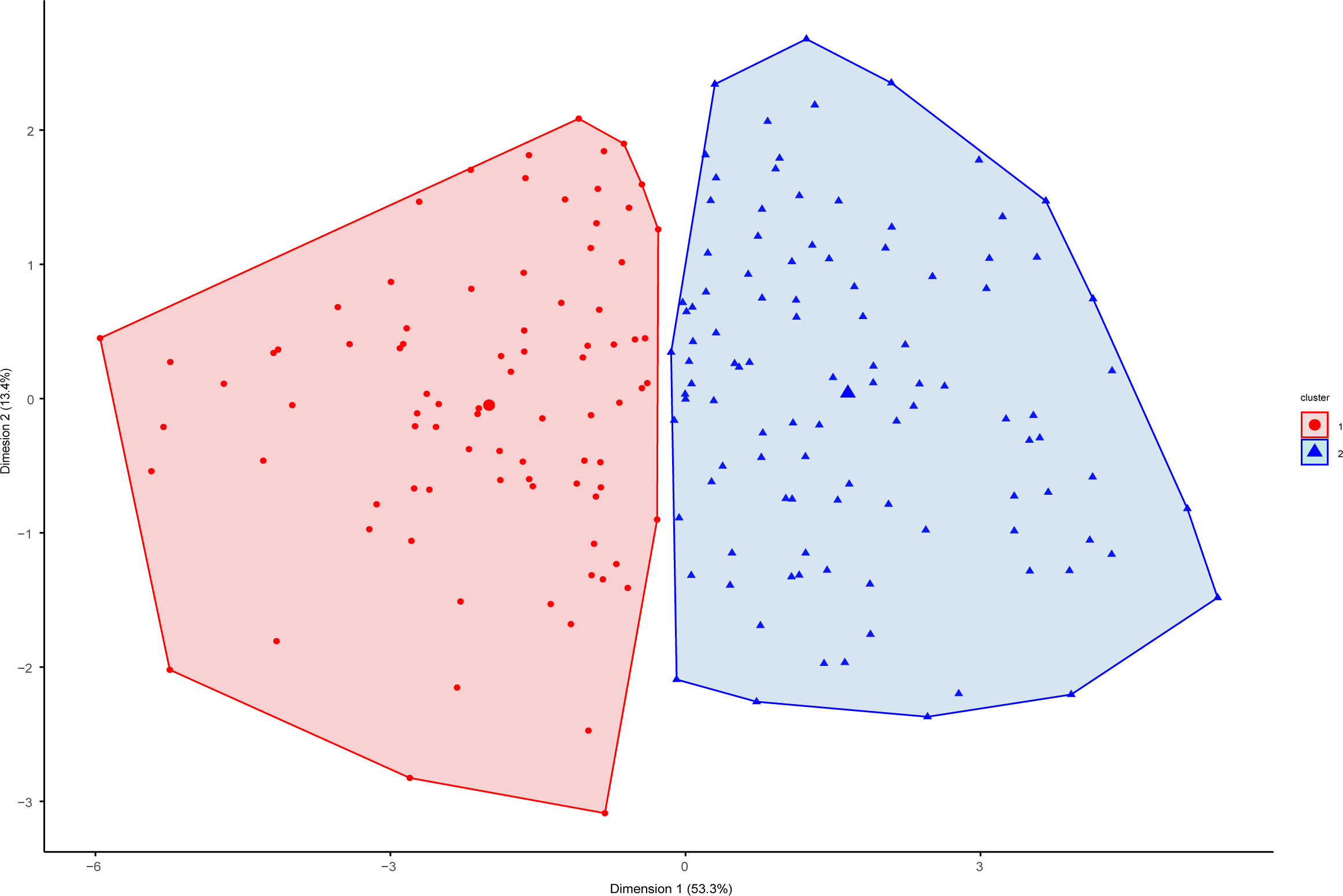
Determination of immune clusters from MCP-counter-based immune deconvoluted cell proportions in the TCGA cohort. Factor map representing two clusters based on hierarchical clustering of principal components. Each dot represents an individual patient sample.

**Extended Data Figure 8.**
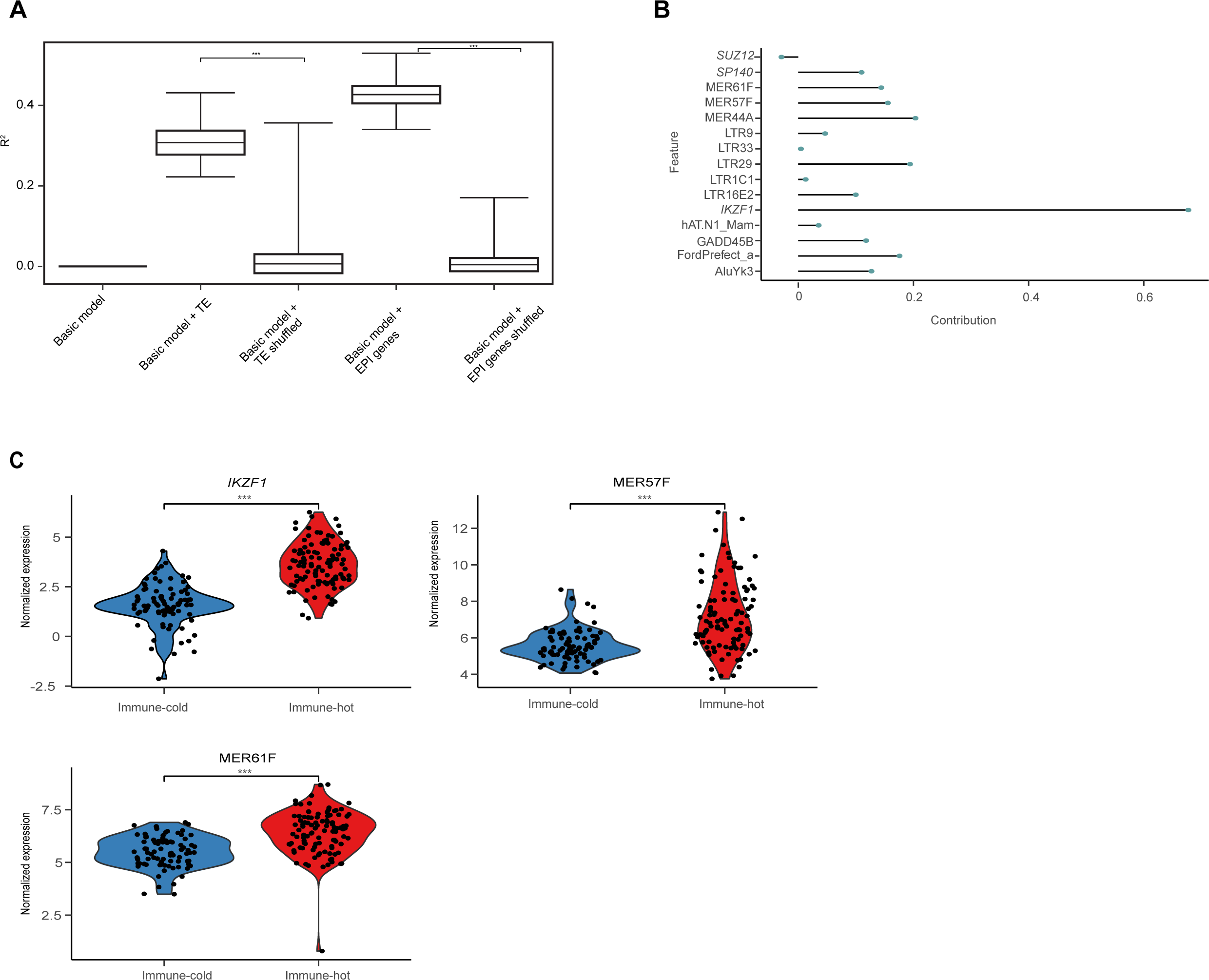
TEs and IKZF1 expression predict immune groups in the TCGA cohort. **A.** Comparison of performances (R2) of 5 lasso logistic regression model models. Each boxplot represents a different model (basic model, bootstraped basic model + TE, and bootstraped basic model + Epigenetic genes (EPI), and error bars represent 95% confidence intervals. Shuffled models for TE and EPI are also shown. Difference between bootstraped and shuffeled model is shown as result of t-test (***, p-value < 2.2e-16). **B.** Contribution of significant features from the TE and epigenetic models (models with the highest R2) represented as non-zero coefficients. Significant TE families and epigenetic genes (italicized) are shown. **C.** Violin plots of normalized expression of transcripts identified as significant features in the regression model in immune-hot and -cold clusters. *** represents p<0.001 as determined by two-sided t-test.

**Extended Data Figure 9.**
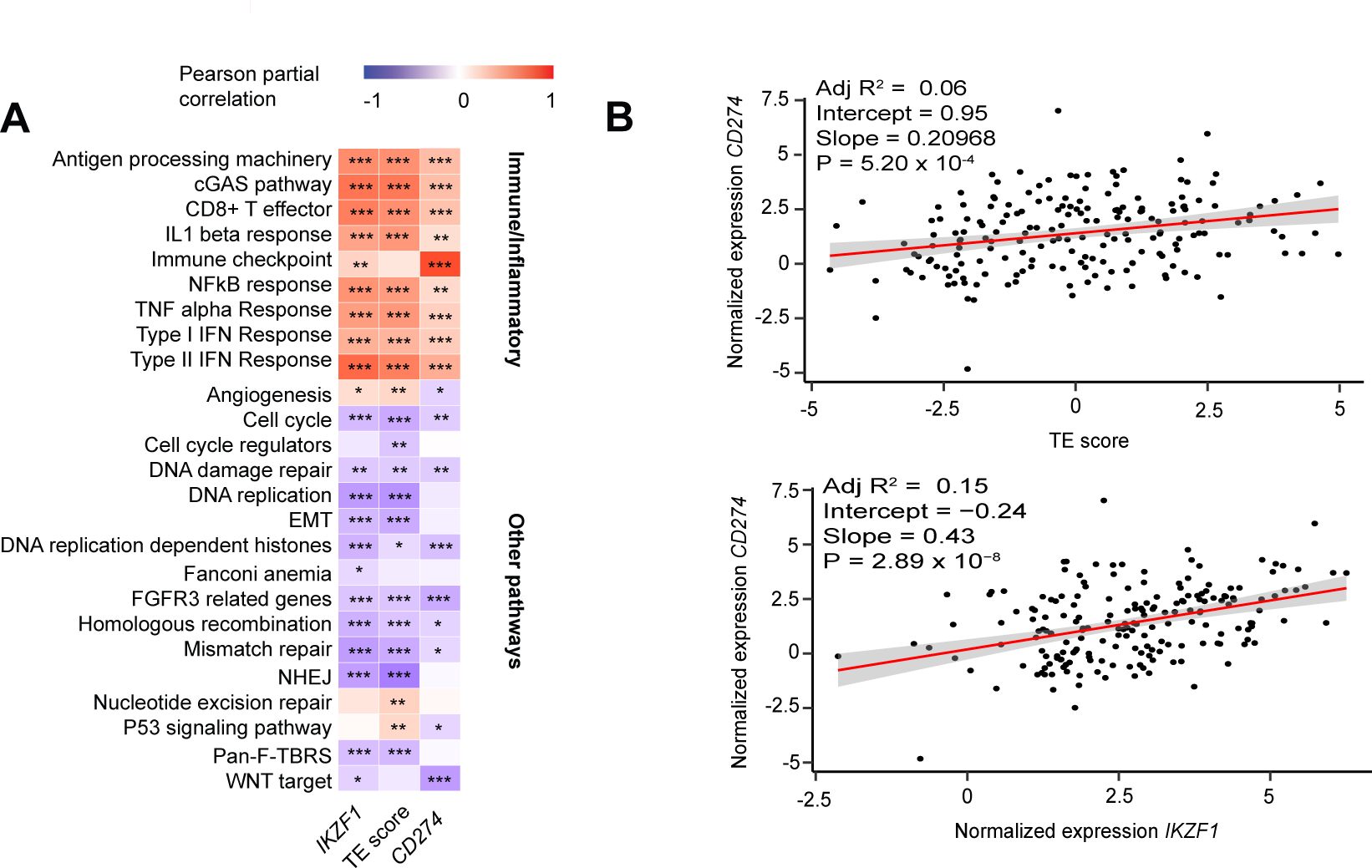
TEs and IKZF1 expression correlate with immune pathway expression in the TCGA cohort. **A.** Heatmap of partial Pearson correlation including batch and histolotgy as covariates. Scale from −1 (inverse correlation, blue), to 1 (positive correlation, red). Asterisks indicate Benjamini-Hochberg-corrected p-values: * p<0.05, ** p<0.01, *** p<0.001. **B.** Correlation between CD274 (PD-L1) gene expression and TE score (top) and IKZF1 expression (bottom).).

## References

1. WHO. Soft tissue and bone tumours, (International Agency for Research on Cancer, Lyon (France), 2020).

2. D’Angelo, S.P., et al. Pilot study of bempegaldesleukin in combination with nivolumab in patients with metastatic sarcoma. Nat Commun 13, 3477 (2022).

3. Cancer Genome Atlas Research, N. Comprehensive and Integrated Genomic Characterization of Adult Soft Tissue Sarcomas. Cell 171, 950-965 e928 (2017).

4. Siegel, R.L., Miller, K.D. & Jemal, A. Cancer statistics, 2018. CA Cancer J Clin 68, 7–30 (2018).

5. von Mehren, M., et al. Soft Tissue Sarcoma, Version 2.2022, NCCN Clinical Practice Guidelines in Oncology. J Natl Compr Canc Netw 20, 815–833 (2022).

6. Seddon, B., et al. Gemcitabine and docetaxel versus doxorubicin as first-line treatment in previously untreated advanced unresectable or metastatic soft-tissue sarcomas (GeDDiS): a randomised controlled phase 3 trial. The Lancet Oncology 18, 1397–1410 (2017).

7. D’Angelo, S.P., et al. Nivolumab with or without ipilimumab treatment for metastatic sarcoma (Alliance A091401): two open-label, non-comparative, randomised, phase 2 trials. The Lancet Oncology 19, 416–426 (2018).

8. Tawbi, H.A., et al. Pembrolizumab in advanced soft-tissue sarcoma and bone sarcoma (SARC028): a multicentre, two-cohort, single-arm, open-label, phase 2 trial. The Lancet Oncology 18, 1493–1501 (2017).

9. Banks, L.B. & D’Angelo, S.P. The Role of Immunotherapy in the Management of Soft Tissue Sarcomas: Current Landscape and Future Outlook. J Natl Compr Canc Netw 20, 834–844 (2022).

10. Carbone, D.P., et al. First-Line Nivolumab in Stage IV or Recurrent Non-Small-Cell Lung Cancer. N Engl J Med 376, 2415–2426 (2017).

11. Hellmann, M.D., et al. Nivolumab plus Ipilimumab in Lung Cancer with a High Tumor Mutational Burden. N Engl J Med 378, 2093–2104 (2018).

12. Rosenberg, J.E., et al. Atezolizumab in patients with locally advanced and metastatic urothelial carcinoma who have progressed following treatment with platinum-based chemotherapy: a single-arm, multicentre, phase 2 trial. The Lancet 387, 1909–1920 (2016).

13. Van Allen, E.M., et al. Genomic correlates of response to CTLA-4 blockade in metastatic melanoma. Science 350, 207–211 (2015).

14. Rizvi, N.A., et al. Cancer immunology. Mutational landscape determines sensitivity to PD-1 blockade in non-small cell lung cancer. Science 348, 124–128 (2015).

15. Samstein, R.M., et al. Tumor mutational load predicts survival after immunotherapy across multiple cancer types. Nat Genet 51, 202–206 (2019).

16. Le, D.T., et al. Mismatch repair deficiency predicts response of solid tumors to PD-1 blockade. Science 357, 409–413 (2017).

17. Nacev, B.A., et al. Clinical sequencing of soft tissue and bone sarcomas delineates diverse genomic landscapes and potential therapeutic targets. Nat Commun 13, 3405 (2022).

18. Italiano, A., et al. Pembrolizumab in soft-tissue sarcomas with tertiary lymphoid structures: a phase 2 PEMBROSARC trial cohort. Nat Med 28, 1199–1206 (2022).

19. Petitprez, F., et al. B cells are associated with survival and immunotherapy response in sarcoma. Nature 577, 556–560 (2020).

20. Allis, C.D., Caparros, M.-L., Jenuwein, T. & Reinberg, D. Epigenetics, (CSH Press, Cold Spring Harbor Laboratory Press, Cold Spring Harbor, New York, 2015).

21. Que, Y., et al. Frequent amplification of HDAC genes and efficacy of HDAC inhibitor chidamide and PD-1 blockade combination in soft tissue sarcoma. J Immunother Cancer 9(2021).

22. Krug, B., et al. Pervasive H3K27 Acetylation Leads to ERV Expression and a Therapeutic Vulnerability in H3K27M Gliomas. Cancer Cell 35, 782–797 e788 (2019).

23. Chiappinelli, K.B., et al. Inhibiting DNA Methylation Causes an Interferon Response in Cancer via dsRNA Including Endogenous Retroviruses. Cell 162, 974–986 (2015).

24. Topper, M.J., et al. Epigenetic Therapy Ties MYC Depletion to Reversing Immune Evasion and Treating Lung Cancer. Cell 171, 1284–1300 e1221 (2017).

25. Sheng, W., et al. LSD1 Ablation Stimulates Anti-tumor Immunity and Enables Checkpoint Blockade. Cell 174, 549–563.e519 (2018).

26. Macfarlan, T.S., et al. Endogenous retroviruses and neighboring genes are coordinately repressed by LSD1/KDM1A. Genes & Development 25, 594–607 (2011).

27. Griffin, G.K., et al. Epigenetic silencing by SETDB1 suppresses tumour intrinsic immunogenicity. Nature (2021).

28. Zhang, S.M., et al. KDM5B promotes immune evasion by recruiting SETDB1 to silence retroelements. Nature 598, 682–687 (2021).

29. Hu, H., et al. Targeting the Atf7ip-Setdb1 Complex Augments Antitumor Immunity by Boosting Tumor Immunogenicity. Cancer Immunol Res 9, 1298–1315 (2021).

30. Burr, M.L., et al. An Evolutionarily Conserved Function of Polycomb Silences the MHC Class I Antigen Presentation Pathway and Enables Immune Evasion in Cancer. Cancer Cell 36, 385–401 e388 (2019).

31. Sturm, G., et al. Comprehensive evaluation of transcriptome-based cell-type quantification methods for immuno-oncology. Bioinformatics 35, i436–i445 (2019).

32. Lê S, J.J., Husson F. FactoMineR: A Package for Multivariate Analysis. Journal of Statistical Software 25, 1–18 (2008).

33. Kelly, C.M., Antonescu, C. R., Bowler, T., Munhoz, R., Chi, P., Dickson, M. A., Gounder, M. M., Keohan, M. L., Movva, S., Dholakia, R., Ahmad, H., Biniakewitz, M., Condy, M., Phelan, H., Callahan, M., Wong, P., Singer, S., Ariyan, C., Bartlett, E. K., Crago, A., Yoon, S., Hwang, S., Erinjeri, J.P., Qin, L.X., Tap, W.D., D’Angelo, S. P. Objective Response Rate Among Patients With Locally Advanced or Metastatic Sarcoma Treated With Talimogene Laherparepvec in Combination With Pembrolizumab: A Phase 2 Clinical Trial. JAMA Oncol, 402–408 (2020).

34. Kelly, C.M., et al. A Phase II Study of Epacadostat and Pembrolizumab in Patients with Advanced Sarcoma. Clin Cancer Res 29, 2043–2051 (2023).

35. Eisenhauer, E.A., et al. New response evaluation criteria in solid tumours: revised RECIST guideline (version 1.1). Eur J Cancer 45, 228–247 (2009).

36. Grundy, E.E., Diab, N. & Chiappinelli, K.B. Transposable element regulation and expression in cancer. FEBS J 289, 1160–1179 (2022).

37. Anwar, S.L., Wulaningsih, W. & Lehmann, U. Transposable Elements in Human Cancer: Causes and Consequences of Deregulation. Int J Mol Sci 18(2017).

38. Kong, Y., et al. Transposable element expression in tumors is associated with immune infiltration and increased antigenicity. Nat Commun 10(2019).

39. Griffin, G.K., et al. Epigenetic silencing by SETDB1 suppresses tumour intrinsic immunogenicity. Nature 595, 309–314 (2021).

40. Churchman, M.L. & Mullighan, C.G. Ikaros: Exploiting and targeting the hematopoietic stem cell niche in B-progenitor acute lymphoblastic leukemia. Exp Hematol 46, 1–8 (2017).

41. Hu, Y., et al. Lineage-specific 3D genome organization is assembled at multiple scales by IKAROS. Cell 186, 5269–5289 e5222 (2023).

42. Soldi, R., et al. The novel reversible LSD1 inhibitor SP-2577 promotes anti-tumor immunity in SWItch/Sucrose-NonFermentable (SWI/SNF) complex mutated ovarian cancer. PLOS ONE 15, e0235705 (2020).

43. Voon, H.P., et al. ATRX Plays a Key Role in Maintaining Silencing at Interstitial Heterochromatic Loci and Imprinted Genes. Cell Rep 11, 405–418 (2015).

44. Stone, M.L., et al. Epigenetic therapy activates type I interferon signaling in murine ovarian cancer to reduce immunosuppression and tumor burden. Proc Natl Acad Sci U S A 114, E10981–E10990 (2017).

45. Chen, J.C., Perez-Lorenzo, R., Saenger, Y.M., Drake, C.G. & Christiano, A.M. IKZF1 Enhances Immune Infiltrate Recruitment in Solid Tumors and Susceptibility to Immunotherapy. Cell Syst 7, 92–103 e104 (2018).

46. Deniz, O., et al. Endogenous retroviruses are a source of enhancers with oncogenic potential in acute myeloid leukaemia. Nat Commun 11, 3506 (2020).

47. Robinson, M.D., McCarthy, D.J. & Smyth, G.K. edgeR: a Bioconductor package for differential expression analysis of digital gene expression data. Bioinformatics 26, 139–140 (2010).

48. Ritchie, M.E., et al. limma powers differential expression analyses for RNA-sequencing and microarray studies. Nucleic Acids Res 43, e47 (2015).

49. Barrero, M.J. Epigenetic Regulation of the Non-Coding Genome: Opportunities for Immuno-Oncology. Epigenomes 4(2020).

50. MIT. Tempo: CCS Research Pipeline for Whole-Genome and Whole-Exome Sequencing (2019).

51. Li, H. Aligning sequence reads, clone sequences and assembly contigs with BWA-MEM. (2013).

52. Smit, A.H., R; Green, P. RepeatMasker Open-4.0. (2013-2015).

53. Karczewski, K.J., et al. The mutational constraint spectrum quantified from variation in 141,456 humans. Nature 581, 434–443 (2020).

54. Shen, R. & Seshan, V.E. FACETS: allele-specific copy number and clonal heterogeneity analysis tool for high-throughput DNA sequencing. Nucleic Acids Res 44, e131 (2016).

55. Friedman, J., Hastie, T. & Tibshirani, R. Regularization Paths for Generalized Linear Models via Coordinate Descent. J Stat Softw 33, 1–22 (2010).

56. Lee, E., Chuang, H.Y., Kim, J.W., Ideker, T. & Lee, D. Inferring pathway activity toward precise disease classification. PLoS Comput Biol 4, e1000217 (2008).

57. Hanzelmann, S., Castelo, R. & Guinney, J. GSVA: gene set variation analysis for microarray and RNA-seq data. BMC Bioinformatics 14, 7 (2013).

